# Effectiveness of data-driven quality improvement on hospitalizations and health outcomes for people with coronary heart disease in primary care (QUEL): a cluster randomised controlled trial with 24-month follow-up

**DOI:** 10.1101/2025.06.12.25329537

**Authors:** Julie Redfern, Nashid Hafiz, Qiang Tu, Andrew Knight, Charlotte Hespe, Clara K Chow, Tom Briffa, Robyn Gallagher, Christopher M Reid, David Hare, Deborah Manandi, Nicholas Zwar, Mark Woodward, Stephen Jan, Emily R Atkins, Tracey-Lea Laba, Elizabeth Halcomb, Laurent Billot, Tim Usherwood, Karice Hyun

## Abstract

**Background:** Using data to drive improved health outcomes offers an healthcare approach that has potential to benefit secondary prevention of coronary heart disease (CHD). The aim of this trial was to test the effectiveness of a data-driven quality improvement program in primary care on cardiovascular hospitalizations, major adverse cardiovascular events (MACE), risk factor profiles and medication prescriptions at 24 months in people with CHD.

**Methods:** Single-blind, cluster randomised controlled trial conducted in 51 Australian primary care practices. Practices having ≥200 adult patients annually with CHD were the units of randomization and people with CHD the units of analysis. Practices were randomised (1:1) to intervention (12-month data-driven quality improvement including benchmarking, monthly reporting, improvement planning) or control (standard care). Primary outcome was proportion of people with CHD who had unplanned CVD hospitalizations at 24-months.

Secondary outcomes were MACE, guideline-recommended medication prescription, risk factor targets, and management planning. Data were extracted from electronic records linked to administrative data and analysed using intention-to-treat log-binomial regression within the framework of generalized estimating equations accounting for clustering of patients within practices. Trial was registered with ANZCTR (ACTRN12619001790134).

**Results:** Between November 2019 and November 2021, 51 primary care practices participated, resulting in a patient cohort of 7864 (4524 from control practices, 3340 from intervention practices). The practices in control and intervention groups were well balanced for practice characteristics, socioeconomic status according to postcode, and median number of patients per practice. Mean age of the patient cohort was 71·9 (±11·8) years, 68% were male and 24% had a prior myocardial infarction. At 24-months, there was no significant in between the control (11·5%) and intervention (10·6%) groups for unplanned CVD hospitalizations (relative risk: 0·91, 95% CI 0·75 to 1·10). Similarly, there was no evidence that secondary outcomes of MACE, clinical measures, or medication prescriptions were different between the groups at 24 months.

**Conclusions:** A primary care, 12-month data-driven quality improvement program did not improve unplanned hospitalizations, MACE or medication prescriptions for people with CHD at 24-month follow-up. Robust evidence for use of a data-driven, collaborative approach to improving care for people with coronary heart disease in primary care remains elusive.

*CONDENSED ABSTRACT:* The QUEL cluster randomized controlled trial tested the effectiveness of data-driven quality improvement on secondary prevention of coronary heart disease in primary care with 24-month follow-up. It is arguably one of the largest and most robust studies evaluating the effectiveness of this type of intervention in primary care or an acute hospital setting. No significant improvement was found in unplanned cardiovascular disease hospitalizations, major adverse cardiovascular events, cardiovascular disease risk factors or medication prescriptions at 12- and 24-months. Evidence for use of a data-driven, collaborative approach to improving care for people with coronary heart disease in primary care remains elusive.

## INTRODUCTION

Cardiovascular disease (CVD), primarily coronary heart disease (CHD), remains the leading cause of death and hospital admissions globally.^1^ With medical advances and improved treatments, CHD death rates are reducing.^2^ Concomitantly, with an aging population, the number of people worldwide living with CHD in need of secondary prevention is increasing).^3^ However, despite widespread recommendations in international guidelines,^4,5,6^ adherence, access, and sustainability of secondary prevention strategies are suboptimal.^7^ Use of evidence-based secondary prevention medications and lifestyle change decline in the initial six months^8^ after an acute nonfatal event and access to cardiac rehabilitation remains poor.^9^ Taken together, there is an escalating need for improved efficiency of and access to effective secondary prevention strategies to reduce hospitalizations and health care costs.^3,7,9^.

The World Heart Federation Roadmap for Secondary Prevention of CVD identifies primary care as offering the most inclusive, equitable, cost-effective, and efficient environment for optimising physical, mental and social well-being.^7^ Collaborative quality improvement for healthcare emerged some 30 years ago and was based on the concept of ‘Breakthrough Series’ developed by The Institute for Healthcare Improvement.^10^ The approach is designed to help organizations improve processes and care through collaborative efforts founded on shared learning from each other and recognized experts to make health improvements.^10^ Breakthrough Series Collaboratives are designed as a short-term (6- to 15-month) learning system that brings together clinical teams to seek improvement in a focused topic area.^10^ Within a collaborative, teams apply quality improvement methods, attend workshops, share learnings, undertake rapid cycle change actions, and are supported by expert facilitators.^10^

Collaborative improvement methodology has been evaluated in a variety of healthcare areas,^11^ including asthma,^12^ chronic heart failure,^13^ and in compliance with healthcare standards.^14^ While such programs have been evaluated, evidence for their effectiveness has predominantly focussed on surrogate endpoints with further robust evidence needed to assess the efficacy of this method for improving care.^11,12,13^ Concomitantly, advances in routine collection and integration of electronic records have enabled more advanced and efficient data availability.^15^ However, current research has not rigorously evaluated the effectiveness of data-driven collaborative quality improvement in primary care in the context of cardiovascular health. The ‘quality improvement in primary care to prevent hospitalizations and improve effectiveness and efficiency of care for people living with CHD’ (QUEL) trial aimed to test whether a 12-month data-driven quality improvement program, delivered in primary care, would reduce the rate of unplanned cardiovascular hospitalizations and improve cardiovascular risk profiles at 24 months among people with CHD. The trial also set out to test the effect on major adverse cardiovascular events (MACE), risk factor profiles, and medication prescriptions at 24 months.

## METHODS

### Study design and participants

QUEL was an investigator-initiated, pragmatic, multicentre, prospective, single-blind cluster randomized controlled trial (cRCT) with 12- and 24-month follow-up. The protocol is published elsewhere.^16^ The trial (ACTRN12619001790134) complied with the CONSORT extension for cRCTs^17^ and the intervention is reported in line with the Template for Intervention Description and Replication (TIDieR).^18^ The trial was conducted at 51 primary care practices across four of the most populous states of Australia (New South Wales, Victoria, Queensland, South Australia) between 2019 and 2022. Participating practices were randomised (1:1) to the intervention (12-month data-driven quality improvement program that included benchmarking, monthly reporting and improvement planning) or control (standard care) regimens. The QUEL trial complied with the Declaration of Helsinki and was approved by the New South Wales Population and Health Services Research Ethics Committee (HREC, HREC/18/CIPHS/44) and appropriate regulatory agencies for linkage of administrative data.

Primary care practices were eligible if they (i) managed ≥ 200 patients year with CHD, (ii) used practice software that was compliant with the data extraction software (PEN Computer Systems) used for collection of clinical data, and (iii) were not participating in a formal quality improvement collaborative program. The patient cohort comprised all eligible patients presenting to participating practices who were (i) 18 years or older, (ii) had a documented diagnosis of CHD in the practice records, and (iii) had visited their general practitioner (GP) at least once in the previous 12 months.

### Randomization and masking

Practices were randomized 1:1 using a computer-generated sequence generated with SAS 9·4 (Proc Surveyselect). Randomization was stratified according to two characteristics - rural versus urban location and size of the practice (≤2 versus > 2 GPs in a practice). The statistician performing randomization and analysis was blinded to practice names and details and only exposed to the practice characteristics to enable stratification. All study data were obtained via linkage between administrative datasets, and no people involved in data linkage or analysis were aware of the group allocation. Practices and the research team members coordinating the intervention (NH, QT, AK, CH) could not be masked to allocation status but did not have access to the data or participate in the analysis.

### Procedure

Eligible practices that agreed to participate entered into a formal agreement, and baseline individual-level clinical data were extracted from practice software prior to randomization. Individual-level clinical data was also extracted from the primary care electronic medical record at 12- and 24-months. Data were securely linked with Australian government administrative data including the Pharmaceutical Benefits Scheme (PBS) for subsidised medication prescriptions, Medical Benefits Scheme (MBS) for health care utilisation, National Death Index for deaths, and relevant state government administrative data for hospitalizations. All data were then combined and analysed in the Secure Unified Research Environment (SURE). Following randomization, practices allocated to the control arm continued with their usual care for all patients attending their practice.

#### Intervention

Practices allocated to the intervention arm participated in a 12-month data-driven quality improvement collaborative based on Breakthrough Series Methodology (Box 1).^10^ The intervention aimed to address the 12 pre-determined key performance indicators relevant to secondary prevention of CHD.^16^ These consisted of measures related to low-density lipoprotein (LDL)-cholesterol, blood pressure (BP), smoking status, medication prescriptions (antiplatelet, statin, angiotensin converting enzyme (ACE) inhibitor or angiotensin II receptor blockers (ARB)), and documentation of a CHD GP Management Plan.^16^ The intervention included participation in learning workshops by practice staff, preparation and submission of rapid improvement (plan-do-study-act, PDSA) cycles using practice data between workshops, and support from local network members to provide technical assistance and provide reminders (Box 1).

##### Box 1

Overview of QUEL Data-driven quality improvement intervention

###### Duration and timeframe

12 months - Nov 2019 to Nov 2020)

###### Virtual orientation session

Practice staff (1-3 team members nominated by the practice) participated in an initial online orientation of 1 hour, which introduced the 12 key performance indicators as targets for improvement, outlined learning workshops, automated data reports, and the PDSA cycles approach to facilitation. Delivered by research team members experienced in collaborative methodology in primary care (AK, CH)

###### Ongoing practice communication

Practice-level SharePoint accounts were established to share the monthly feedback reports, submit PDSA cycles and to access trial resources.

###### Learning workshops

- One full day face-to-face workshop (held in Sydney Australia) to introduce change principles, evidence for indicators, understanding practice-level population, creating an automated recall system, and practices shared real-time experiences on how they implement activities to drive improvement
- Five virtual workshops (one hour each) summarising monthly aggregate reports, PDSA cycles, understanding benchmarking of indicators and enhancing CHD management through care planning and shared learning with other participating practices, shared planning of sustained improvement
- Workshops were delivered by primary care, quality improvement and CVD experts

###### Activity periods (between the learning workshops)

- Practice data were auto-extracted monthly from the electronic medical records and uploaded to SharePoint for practices to use in formulation and monitoring their PDSA cycles.
- Aggregate (whole of collaborative) reports were also provided to enable practices to benchmark their progress, compare themselves to the performance of others, and to provide motivation. For example, if there were low numbers of patients on statin medications at a practice compared to others.

###### Support and facilitation

- Staff from local primary health networks or a QUEL team member (NH, QT) provided monthly support and facilitation to practices between learning workshops including telephone support to assist with technical issues and remind practices about PDSA submission and improvement strategies.
- QUEL, Quality improvement in primary care to prevent hospitalizations and improve Effectiveness and efficiency of care for people Living with CHD; PDSA, plan-do-study-act; CHD, coronary heart disease; CVD, cardiovascular disease

### Outcomes

The primary outcome was the proportion of patients with at least one unplanned CVD hospitalization within 24-months from baseline data collection. For this trial, CVD was defined as any condition involving the heart, brain or peripheral blood vessels and included CHD (such as angina and myocardial infarction), cerebrovascular disease (such as stroke), peripheral arterial disease, heart failure, and atrial fibrillation.^19^ (Primary outcome data were collected via probability matched and privacy preserved linkage of the practice cohorts with individual-level state/territory government administrative data for hospital admissions (Supplementary Table 1).

Secondary outcomes were:

1. Proportion of patients with MACE that included CHD (unstable angina or myocardial infarction), stroke or CVD death (fatal and non-fatal) collected via probability matched and privacy preserved linkage of the practice cohort with individual-level state government hospital readmissions and federal government death index administrative data at 24-months (see Supplementary Table 1 for codes).
2. Proportion of patients who received guideline-recommended medicines (i.e. anti-platelet, statin and ACE inhibitors or ARB collected via linkage of the practice cohort with individual-level federal government administrative data for pharmaceutical prescriptions in Australia (via the PBS) at 24-months.
3. Proportion of patients with a prepared General Practice Management Plan (MBS Item #721), prepared Team Care Arrangement (MBS Item #723), or an associated review (MBS Item #732) collected via linkage of the practice data (within preceding five months) with individual-level federal government administrative data at 24-months. These plans are funded by the Australian Medicare system to support care planning and review in primary care.
4. Proportion of patients not achieving national targets for CVD risk factors (LDL≥2mmol/l, systolic BP (SBP) >130mmHg, diastolic BP (DBP) >80 mmHg, and current smoker, collected via primary care clinical data extraction at 24 months.
5. Primary and secondary outcomes at 12-months from baseline.

Engagement with the intervention was summarised through an end of intervention user survey combined with submission of PDSA cycles and attendance at learning workshops.^20^ The user survey included Likert questions about usefulness and satisfaction, as well as free-text questions exploring barriers and enablers to engagement. Practice team members including general practitioners, practice managers, or nurses; from intervention practices who were actively involved in implementing data-driven improvements within their practices were invited to complete the survey. De-identified survey responses were entered into an online database and analysed using descriptive statistics. Free text responses were coded using thematic analysis.

### Statistical Analysis

The target number of participating practices was 50. The target individual sample size was 6050 (3025 per group) with an average cluster size of 121 patients per practice. Using a 5% two-sided test, this was estimated to provide 80% power to detect a relative risk of 0·75. This calculation assumed a control group readmission rate of 35% based on an Australian cohort study reporting an atherothrombotic disease readmission rate of 35% at two years for patients with CHD.^21^ An intra-class correlation coefficient of 0·05 was assumed, based on data from two cross-sectional studies in Australian primary care.^22^

Analysis was conducted at the individual patient level following the intention-to-treat principle with data analyzed according to randomization group. Baseline patient characteristics, cardiovascular risk, and treatment usage were summarized using frequency and percentage for categorical variables and mean and standard deviation for continuous variables. Primary analysis was conducted using the log-binomial regression within the framework of generalized estimating equations with an exchangeable correlation to account for the clustering of patients within general practices. The effect of the intervention is presented as a relative risk (RR) and 95% confidence interval (CI). A planned sensitivity analysis of time to unplanned CVD admission was performed using a Fine-Gray competing risks model with robust sandwich covariance matrix estimate to account for the competing risk of death and to provide valid standard errors that are robust to possible misspecification of the assumed correlation structure, including potential clustering within general practices. Sensitivity analysis results are reported as sub-distribution hazard ratio and 95% CI. An exploratory subgroup analyses was also undertaken to examine the effects of sex on the primary outcome. Secondary outcomes were analysed using the log-binomial regression within the framework of generalized estimating equations with an exchangeable correlation, reported as RR and 95% CI. All analyses were performed using a two-sided significance level of 0·05 in SAS version 9·4 (SAS Institute Inc., Cary, NC, USA).

### Data sharing statement

Due to constraints related to ethical approval for the QUEL trial, participant level data cannot be shared outside the SURE.

## RESULTS

A total of 51 primary care practices participated in the QUEL trial with baseline data collected on Nov 10 2019 and final extraction on Dec 10 2021. This resulted in an individual level patient cohort of 7864; 4524 from 25 control practices and 3340 from 26 intervention practices (Figure 1). All practices provided clinical data for linkage so there was no loss to follow-up at 12-or 24-months for the primary outcome. Primary care clinical records (LDL, SBP, smoking status, BMI) could not be collected at 12- and 24-month follow-up for all patients due to a change in the primary care electronic medical record software since baseline (Table 2). However, all data were obtained for the primary outcome, MACE, medication prescriptions (PBS), and General Practice Management Plan (MBS).

**Figure 1:**
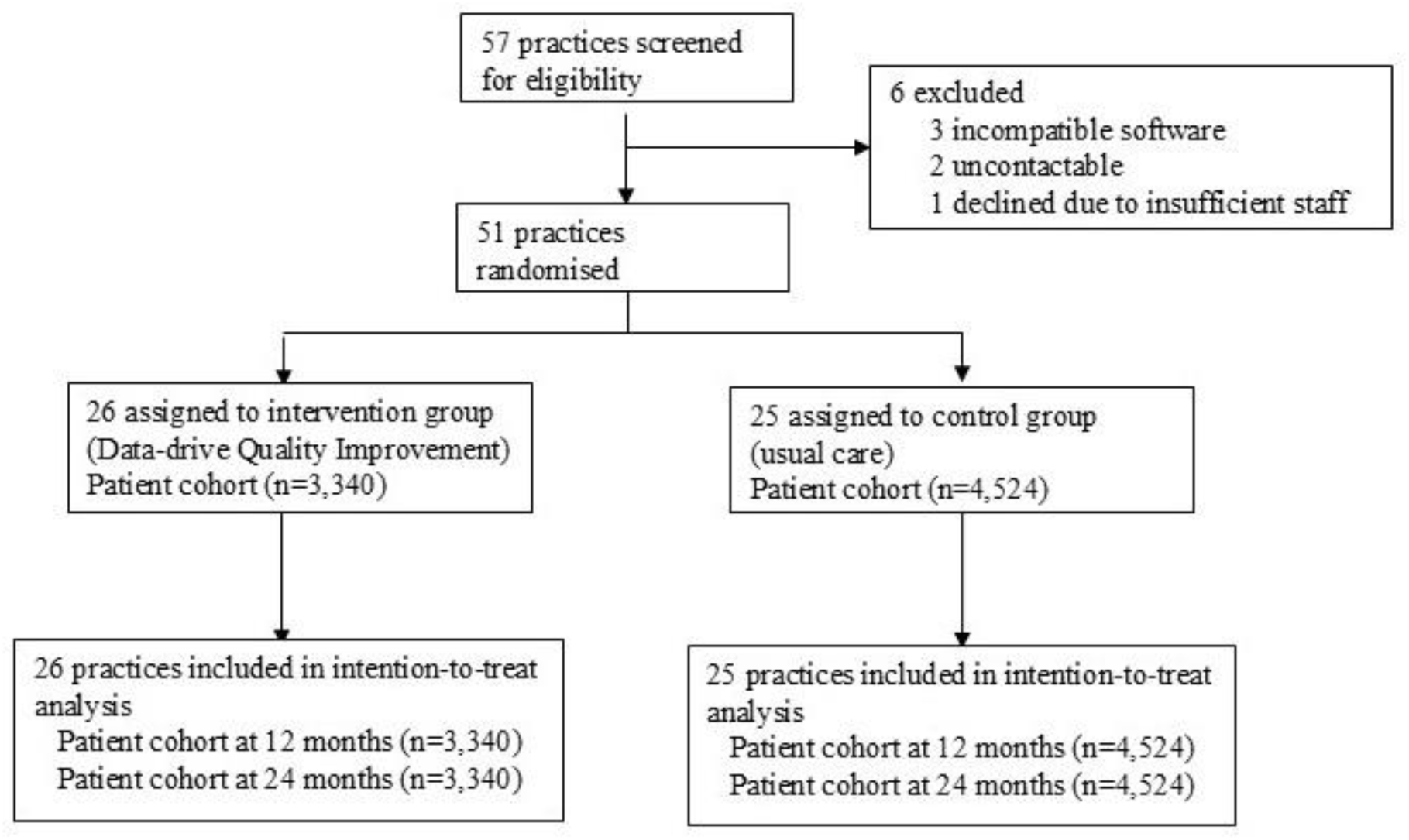
QUEL Trial flow diagram

The practices in control and intervention groups were well balanced for practice characteristics, socioeconomic status according to postcode, and median [inter-quartile interval (IQI)] number of patients per practice (187 [129, 470] versus 187 [110, 318]), and number of GPs working at each practice (4 [2, 9) versus 7 [3, 10]) for control and intervention groups respectively. In terms of the patient cohort, the groups were well balanced at baseline in terms of practice characteristics, patient demographics and clinical measures but there were more patients with CHD attending control practices (Table 1).

**Table 1:**
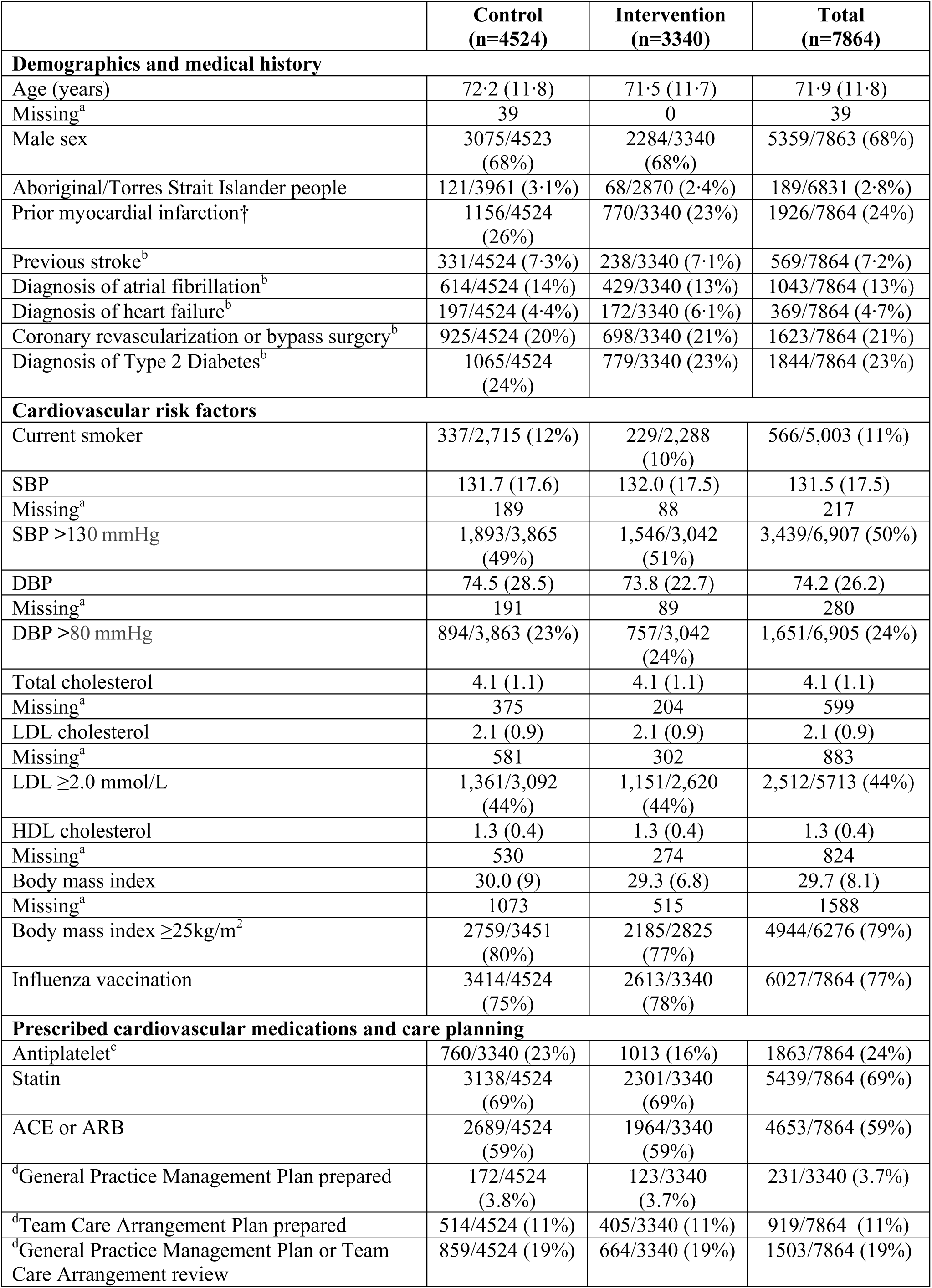
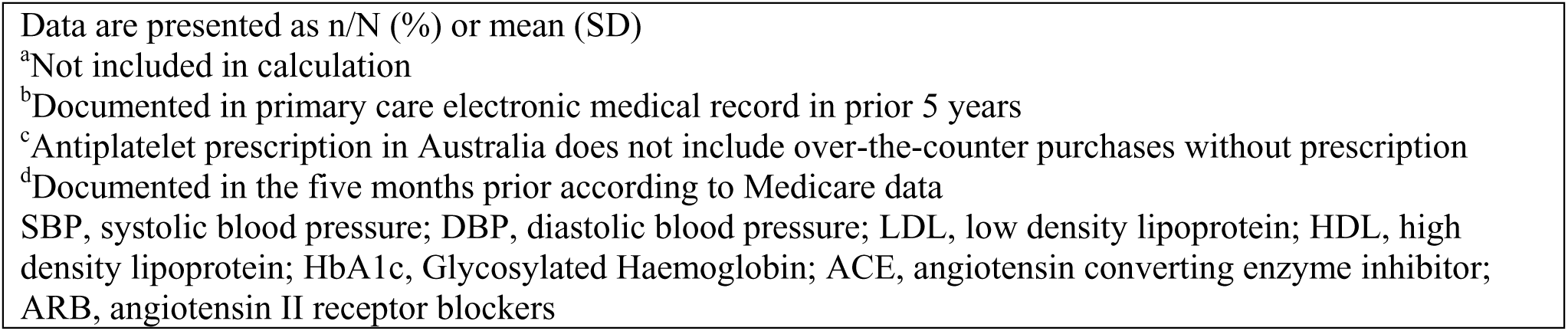
Baseline demographics and characteristics.

| **Table 1: Baseline demographics and characteristics** |  |  |  |
| --- | --- | --- | --- |
|  | **Control**<br>**(n=4524)** | **Intervention**<br>**(n=3340)** | **Total**<br>**(n=7864)** |
| **Demographics and medical history** |  |  |  |
| Age (years) | 72.2 (11.8) | 71.5 (11.7) | 71.9 (11.8) |
| Missing<sup>a</sup> | 39 | 0 | 39 |
| Male sex | 3075/4523<br>(68%) | 2284/3340<br>(68%) | 5359/7863 (68%) |
| Aboriginal/Torres Strait Islander people | 121/3961 (3.1%) | 68/2870 (2.4%) | 189/6831 (2.8%) |
| Prior myocardial infarction† | 1156/4524<br>(26%) | 770/3340 (23%) | 1926/7864 (24%) |
| Previous stroke<sup>b</sup> | 331/4524 (7.3%) | 238/3340 (7.1%) | 569/7864 (7.2%) |
| Diagnosis of atrial fibrillation<sup>b</sup> | 614/4524 (14%) | 429/3340 (13%) | 1043/7864 (13%) |
| Diagnosis of heart failure<sup>b</sup> | 197/4524 (4.4%) | 172/3340 (6.1%) | 369/7864 (4.7%) |
| Coronary revascularization or bypass surgery<sup>b</sup> | 925/4524 (20%) | 698/3340 (21%) | 1623/7864 (21%) |
| Diagnosis of Type 2 Diabetes<sup>b</sup> | 1065/4524<br>(24%) | 779/3340 (23%) | 1844/7864 (23%) |
| **Cardiovascular risk factors** |  |  |  |
| Current smoker | 337/2,715 (12%) | 229/2,288<br>(10%) | 566/5,003 (11%) |
| SBP | 131.7 (17.6) | 132.0 (17.5) | 131.5 (17.5) |
| Missing<sup>a</sup> | 189 | 88 | 217 |
| SBP >130 mmHg | 1,893/3,865<br>(49%) | 1,546/3,042<br>(51%) | 3,439/6,907 (50%) |
| DBP | 74.5 (28.5) | 73.8 (22.7) | 74.2 (26.2) |
| Missing<sup>a</sup> | 191 | 89 | 280 |
| DBP >80 mmHg | 894/3,863 (23%) | 757/3,042<br>(24%) | 1,651/6,905 (24%) |
| Total cholesterol | 4.1 (1.1) | 4.1 (1.1) | 4.1 (1.1) |
| Missing<sup>a</sup> | 375 | 204 | 599 |
| LDL cholesterol | 2.1 (0.9) | 2.1 (0.9) | 2.1 (0.9) |
| Missing<sup>a</sup> | 581 | 302 | 883 |
| LDL ≥2.0 mmol/L | 1,361/3,092<br>(44%) | 1,151/2,620<br>(44%) | 2,512/5713 (44%) |
| HDL cholesterol | 1.3 (0.4) | 1.3 (0.4) | 1.3 (0.4) |
| Missing<sup>a</sup> | 530 | 274 | 824 |
| Body mass index | 30.0 (9) | 29.3 (6.8) | 29.7 (8.1) |
| Missing<sup>a</sup> | 1073 | 515 | 1588 |
| Body mass index ≥25kg/m<sup>2</sup> | 2759/3451<br>(80%) | 2185/2825<br>(77%) | 4944/6276 (79%) |
| Influenza vaccination | 3414/4524<br>(75%) | 2613/3340<br>(78%) | 6027/7864 (77%) |
| **Prescribed cardiovascular medications and care planning** |  |  |  |
| Antiplatelet<sup>c</sup> | 760/3340 (23%) | 1013 (16%) | 1863/7864 (24%) |
| Statin | 3138/4524<br>(69%) | 2301/3340<br>(69%) | 5439/7864 (69%) |
| ACE or ARB | 2689/4524<br>(59%) | 1964/3340<br>(59%) | 4653/7864 (59%) |
| <sup>d</sup>General Practice Management Plan prepared | 172/4524<br>(3.8%) | 123/3340<br>(3.7%) | 231/3340 (3.7%) |
| <sup>d</sup>Team Care Arrangement Plan prepared | 514/4524 (11%) | 405/3340 (11%) | 919/7864 (11%) |
| <sup>d</sup>General Practice Management Plan or Team Care Arrangement review | 859/4524 (19%) | 664/3340 (19%) | 1503/7864 (19%) |

Data are presented as n/N (%) or mean (SD)
<sup>a</sup>Not included in calculation
<sup>b</sup>Documented in primary care electronic medical record in prior 5 years
<sup>c</sup>Antiplatelet prescription in Australia does not include over-the-counter purchases without prescription
<sup>d</sup>Documented in the five months prior according to Medicare data
SBP, systolic blood pressure; DBP, diastolic blood pressure; LDL, low density lipoprotein; HDL, high density lipoprotein; HbA1c, Glycosylated Haemoglobin; ACE, angiotensin converting enzyme inhibitor; ARB, angiotensin II receptor blockers

**Table 2:** Analysis of primary and secondary outcomes

| **Table 2: Analysis of primary and secondary outcomes** |  |  |  |
| --- | --- | --- | --- |
|  | **Control**<br>**(n=4524)** | **Intervention**<br>**(n=3340)** | **Adjusted**<br>**Relative risk**<br>**(95% CI)** |
| Unplanned CVD hospitalization | 520/4524 (11.5%) | 355/3340 (10.6%) | 0.91 (0.75 to 1.10) |
| Unplanned CVD hospitalization 12-months | 306/4524 (6.8%) | 207/3340 (6.2%) | 0.90 (0.70 to 1.15) |
| Major adverse cardiovascular event | 288/4524 (6.4) | 185/3340 (5.5) | 0.81 (0.61 to 1.07) |
| Major adverse cardiovascular event 12-months | 163/4524 (3.6%) | 115/3340 (3.4%) | 0.89 (0.65 to 1.23) |
| Antiplatelet prescription<sup>a</sup> | 809/4524 (17.9%) | 568/3340 (17.0%) | 0.94 (0.79 to 1.13) |
| Statin prescription | 2946/4524 (65.1%) | 2225/3340 (66.6%) | 1.03 (0.97 to 1.09) |
| ACE or ARB prescription | 2468/4524 (54.6%) | 1824/3340 (54.6%) | 1.00 (0.93 - 1.07) |
| General Practice Management Plan prepared<sup>b</sup> | 114/4524 (2.5%) | 96/3340 (2.9%) | 0.85 (0.45 to 1.61) |
| Team Care Arrangement Plan prepared<sup>b</sup> | 389/4524 (8.6%) | 115/3340 (3.4%) | 0.99 (0.72 - 1.34) |
| General Practice Management Plan or Team Care Arrangement review<sup>b</sup> | 673/4524 (14.9%) | 472/3340 (14.1%) | 1.02 (0.64 to 1.63) |
| LDL≥2.0 mmol/L | 1018/2665 (38.2%) | 849/2225 (38.2%) | 0.99 (0.86 to 1.13) |
| SBP >130 mmHg | 1804/3347 (53.9%) | 1298/2507 (51.8%) | 0.97 (0.87 to 1.09) |
| DBP >80 mmHg | 815/3347 (24.4%) | 593/2507 (23.7%) | 0.97 (0.81 to 1.15) |
| Current smoker | 135/1041 (13.0%) | 115/787 (14.6%) | 0.96 (0.57 to 1.59) |
| Data are presented as n/N (%) at 24 months follow-up unless otherwise stated |  |  |  |
| <sup>a</sup>Antiplatelet prescription in Australia does not include over-the-counter purchases without prescription |  |  |  |
| <sup>b</sup>Documented in the five months prior according to Australian Medicare data |  |  |  |
| CVD, cardiovascular disease; ACE, angiotensin converting enzyme inhibitor; ARB, angiotensin II receptor blocker; LDL, low density lipoprotein cholesterol; SBP, systolic blood pressure; DBP, diastolic blood pressure |  |  |  |

For the primary outcome, the rate of unplanned CVD hospitalizations at 24 months was not significantly different between the control (11·5%) and intervention (10·6%) groups (relative risk: 0·91, 95% CI 0·75 to 1·10) (Table 2). There were a total of 432 deaths with no statistically significant difference between control (270/4524; 6·0%) and intervention groups (162/3340; 4·9%) at 24 months. Similarly, there were no significant differences in secondary outcomes between the intervention and control groups (Table 2).

The planned sensitivity analysis to assess the effect of the intervention compared to the control on time to unplanned cardiovascular hospitalization accounting for competing risk of death revealed the difference remained non-significant (sub-distribution hazard ratio: 0·92 (95% CI: 0·73 to 1·16). Subgroup analysis also found no difference in the effect of the intervention on the primary outcome compared to control between sexes (Supplementary Table 2).

Of the practices allocated to intervention, 85% (22/26) attended three or more of the learning workshops and 69% (18/26) submitted at least one PDSA cycle. When asked about satisfaction with the learning workshops, 94% of participating practices rated Learning Workshop One (in-person) as highly satisfactory and 76% rated Learning Workshop Six (virtual) as highly satisfactory. Further, 90% of practices reported that they were “well informed about the objectives of the workshops,” and 85% reported that they felt able to use what they learned in the workshops.

## DISCUSSION

QUEL was an investigator-initiated cluster randomised clinical trial that tested the effectiveness of data-driven quality improvement to strengthen secondary prevention of CHD in primary care. It is arguably one of the largest and most robust studies evaluating the effectiveness of this type of intervention in primary care or an acute hospital setting. No statistically significant improvement was found in unplanned CVD hospitalizations, MACE, CVD risk factors or medication prescriptions at 12- and 24-months. The data-driven and collaborative approach aimed to promote shared learning and quality improvement among participating practices and the majority of intervention practices participated in workshops and submitted PDSAs.

The QUEL intervention included all the standard aspects of a Breakthrough Series collaborative with a similar intervention duration, number of sites and length of follow-up to other studies evaluating similar interventions.^10^ Despite many previous studies evaluating collaborative quality improvement have reported positive process and qualitative findings; the majority have had weak designs and surrogate endpoints.^11^ Three relevant cRCTs have been conducted in primary care; two had robust designs and analysis and tested the intervention in improving asthma care^12^ and colorectal screening;^23^ another tested the intervention for diabetes care in a low resource setting.^24^ The asthma care and colorectal screening trials both found no significant effect of the intervention on their primary (asthma action plans and colorectal screening rates respectively)^12,23^ or secondary outcomes, including hospitalizations for the asthma trial.^12^ The trial evaluating diabetes care did show significant improvement in the intervention group, and not the control group, but analyses were not adjusted and did not directly compare groups.^24^ Another relevant, well-designed cRCT evaluated collaborative quality improvement for heart failure care in a hospital setting and found no significant effect on heart failure processes of care or workforce.^13^ The QUEL trial included a large patient cohort and robust endpoints such as hospitalizations and MACE and taken together, it is argued that QUEL trial adds to the body of evidence suggesting data-driven and collaborative quality improvement may not be effective in improving clinical outcomes for people with complex health conditions in primary care.

The QUEL trial provides insights for future decentralised clinical trials linking primary care data with other administrative health data. The Food and Drug Administration describe defined decentralized clinical trials as those where some or all trial-related activities occur at locations other than traditional clinical trial sites (e.g., the participant’s home, mobile research units, or local health care facilities).^25^ This design is relevant to QUEL given the remote data collection and data-driven nature of the intervention with virtual and telephone support to practices. QUEL offers insight into some of the strengths and weaknesses of such an approach. Strengths include the streamlined recruitment of a large cohort without individualized in-person visits (improved efficiency and reduced resources) and the minimal loss to follow-up.^26,27^ Further, QUEL demonstrated how Australian primary care data can be linked with jurisdictional and national data within the context of a clinical trial. This issue is particularly interesting in Australia where hospitalization data is collected by state/territory governments yet the national government collects deaths and medication prescription data. Efforts to overcome challenges with sharing and linking individual-level patient data, as was the case for QUEL, could improve efficiency of large-scale trials.

There are limitations associated with the QUEL trial. The collection of data via linkage is efficient and pragmatic but also means the quality of reported outcomes is dependent on the quality of data entered at the point of care. This could be an issue for the primary care data if practice-level data collection is not up-to-date. Administrative data for hospitalizations in Australia is collected at a state-based level and QUEL included only four of Australians seven states/territories. This potentially means if a person regularly attended a practice in an included state but travelled to and was admitted to hospital in a non-participating state, their admission may not have been captured. However, the likelihood of this occurring is extremely low. It is possible, the COVID-19 pandemic negatively impacted the ability of intervention practices to prioritize data-driven quality improvement for secondary prevention of CHD. The pandemic may also have impacted hospitalization rates given the observed rate of the primary outcome being much lower than anticipated.

## Conclusion

A 12-month data-driven quality improvement primary care did not result in improvement in unplanned hospitalizations, MACE or medication prescriptions for people with CHD at 24-months. The trial was arguably one of the largest and most robust studies evaluating the effectiveness of this type of intervention in primary care or an acute hospital setting and does add to the growing body of studies showing a lack of evidence. The nature of the study design enabled to the trial to have a large sample size with robust outcomes in an efficient way and provides insight for future trials. Use of a collaborative approach to improving care for people with CHD and using routinely collected primary care data offers opportunity but evidence remains elusive.

## SOURCES OF FUNDING

Funding for this trial was provided by a National Health and Medical Research Council (NHMRC) Partnership Project Grant (Award Grant Number: GNT1140807), Canberra ACT. JR, CKC and KH are funded by a NHMRC Investigator Grants (GNT2007946, GNTP1195326 and GNT1196724 respectively) Canberra, ACT. Additional in-kind and cash support from the following partner organisations; Amgen (Sydney, NSW), Austin Health (Melbourne Victoria), Australian Cardiovascular Health and Rehabilitation Association (Chittaway Bay, NSW), Australian Commission on Safety and Quality in Health Care (Sydney, NSW), Australian Primary Health Care Nurses Association, Brisbane South Primary Health Network (Brisbane Qld, Fairfield General Practice Unit, Heart Support Australia (Phillip, ACT), Improvement Foundation (Adelaide, SA), Inala Primary Care (Inala, Queensland), National Heart Foundation of Australia (Melbourne, Victoria), Nepean Blue Mountains Primary Health Network (Penrith, NSW), Royal Australian College of General Practitioners (North Sydney, NSW), Sanofi (provided cash support via the Externally Sponsored Collaboration Pathway, Macquarie Park, NSW), South Western Sydney Primary Health Network, The George Institute for Global Health (cash support) and University of Melbourne (Melbourne, Victoria).

## DISCLOSURES

We declare no directly relevant competing interests. This was an investigator-initiated trial and the funders had no role in trial design, data collection, analysis, interpretation, or manuscript writing.

## CONTRIBUTORS

JR, CKC, TB, RG, CR, DH, NZ, MW, SJ, EA, TL, EH, LB, TU, KH developed the trial design and scientific oversight. JR, NH, AK, CH, EA, TL, TU led finalisation of key performance indicators. JR, NH, AK, CH, DM supported and contributed to learning workshops and oversight of PDSA cycles. KH completed the analysis. All authors have reviewed, contributed to, and approved the manuscript.

## Data Availability

Due to ethics requirements data cannot be made publicly available.

## Notes

### Competing Interest Statement

The authors have declared no competing interest.

### Clinical Trial

ANZCTR (ACTRN12619001790134)

### Clinical Protocols

https://bmcprimcare.biomedcentral.com/articles/10.1186/s12875-020-01105-0

### Author Declarations

The QUEL trial complied with the Declaration of Helsinki and was approved by the New South Wales Population and Health Services Research Ethics Committee (HREC, HREC/18/CIPHS/44) and appropriate regulatory agencies for linkage of administrative data.

